# A meta-analysis of the genome-wide association studies on two genetically correlated phenotypes (self-reported headache and self-reported migraine) identifies four new risk loci for headaches (N=397,385)

**DOI:** 10.1101/2021.09.15.21263668

**Authors:** Weihua Meng, Parminder S Reel, Charvi Nangia, Aravind Lathika Rajendrakumar, Harry L Hebert, Mark J Adams, Hua Zheng, Zen Haut Lu, 23andMe Research Team, Debashree Ray, Lesley A Colvin, Colin NA Palmer, Andrew McIntosh, Blair H Smith

## Abstract

Headache is one of the commonest complaints that doctors need to address in clinical settings. The genetic mechanisms of different types of headache are not well understood. In this study, we performed a meta-analysis of genome-wide association studies (GWAS) on the self-reported headache phenotype from the UK Biobank cohort and the self-reported migraine phenotype from the 23andMe resource using the metaUSAT for genetically correlated phenotypes (N=397,385). We identified 38 loci for headaches, of which 34 loci have been reported before and 4 loci were newly identified. The *LRP1-STAT6-SDR9C7* region in chromosome 12 was the most significantly associated locus with a leading *P* value of 1.24 × 10^−62^ of rs11172113. The *ONECUT2* gene locus in chromosome 18 was the strongest signal among the 4 new loci with a *P* value of 1.29 × 10^−9^ of rs673939. Our study demonstrated that the genetically correlated phenotypes of self-reported headache and self-reported migraine can be meta-analysed together in theory and in practice to boost study power to identify more new variants for headaches. This study has paved way for a large GWAS meta-analysis study involving cohorts of different, though genetically correlated headache phenotypes.

## Background

Headache is one of the commonest symptoms that present to clinicians in general practice or in specialist neurology clinics. (1) Its lifetime prevalence in individuals is as high as 93%. (2) Globally, around 46% of the adult population suffers with an active headache disorder. (3) According to the current definitions of the International Headache Society, headaches can be classified into three categories: 1. primary headaches (including migraine, tension-type headache, and trigeminal autonomic cephalalgias); 2. secondary headaches (including headaches attributed to other disorders such as trauma, infection); 3. painful cranial neuropathies, other facial pain and other headaches. (4)

Among all types of headache, tension-type headache is the commonest form, causing over 40% of all headaches in the general population, while migraine is the most disabling type at population level, with a prevalence of around 10% of all headaches. (5) It is important to note that an individual can experience more than one type of headache at the same time. (1)

According to the Global Burden of Diseases 2019 study, headache disorders represent the 14th leading cause of disability-adjusted life years (DALYs) when considering all ages and both genders. (6) As the definition of the headache disorders in this study only included migraine and tension type headache, it is reasonable to assume that the real impact of all types of headaches in the world is much more significant. It was estimated in 2003 that migraine alone cost the UK over £2bn a year. (7) Migraine is still one of the leading causes of disability among over 300 diseases. (8)

It has been confirmed that headaches such as migraine are heritable. The single nucleotide polymorphism (SNP) based-heritabilities of migraine and self-reported headache were 0.15 and 0.21 in Caucasians, respectively. (9,10) Genome-wide association studies (GWAS) have revealed that there are significant genetic components contributing to migraine. (11-15) A GWAS meta-analysis paper consisting of 22 cohorts by Gormley et al identified 38 genetic loci for migraine. (9) Our study based on the UK Biobank resource also revealed 28 risk loci for self-reported headache, of which 14 loci had been previously identified by Gormley et al and 14 loci were newly reported. (10)

Recently, researchers have been encouraged to perform GWAS meta-analysis on genetically correlated phenotypes, due to the increasing recognition of pleiotropy in GWAS, to boost study power to detect more genetic components. (16) Pleiotropy refers to the phenomenon where a genetic variant or a gene has non-zero effect on multiple phenotypic traits, and can contribute to genetic correlations among these traits. (17) Successful examples of recent GWAS meta-analysis studies on genetically correlated phenotypes have included education and intelligence as well as different hypertension phenotypes. (18,19)

In a previous study, we reported that the self-reported headache phenotype from the UK Biobank and the self-reported migraine phenotype from the 23andMe were genetically correlated, with a high correlation value of 0.72. (20) Therefore, we aimed to perform a joint GWAS meta-analysis study of these two different but highly genetically correlated phenotypes with a view to replicating previously identified genetic associations and identifying new associations arising from the increased power of this approach.

## Methods

### Cohorts’ information

The two sets of GWAS summary statistics used in this study were from the GWAS on self-reported headache based on the UK Biobank cohort and the GWAS on self-reported migraine provided by the 23andMe. (9,10)

The definitions of self-reported headache (UK Biobank) were: cases (N= 74,461), defined as those who self-reported headache symptoms affecting daily lives within last month using the UK Biobank online questionnaire; controls (N= 149,312), defined as those who did not have any pain affecting daily lives within last month. The corresponding GWAS analysis was performed using a linear mixed model adjusting for age, sex, nine population principal components, genotyping arrays, and assessment centers (10). The dataset contains 9,304,965 SNPs (minor allele frequency > 0.005, imputation score > 0.1).

The definitions of self-reported migraine (23andMe) were: cases (N=30,465), defined as those who self-reported a migraine history (diagnosed by doctors or self-diagnosing) using the 23andMe online questionnaire; controls (N=143,147), those who self-reported having no migraine. The corresponding GWAS was performed using a linear mixed model adjusting for age, sex, and five population principal components (9). The dataset contains 19,023,436 SNPs (containing minor allele frequency and imputation score information for all SNPs).

All of the participants in both GWAS were of European descent. In addition, as the UK Biobank cohort only recruited within the UK, while the 23andMe mainly recruited from the USA, there was little sample overlap between the two cohorts (linkage disequilibrium score regression intercept = 0.009). (20) The detailed cohorts’ information and the statistical methods of the two GWAS can be found in the original papers. (9,10).

### The preprocessing of the GWAS summary statistics

SNPs in both datasets were coded in a forward direction and according to the GRCh37 genome build. In total, 8,500,802 SNPs with minor allele frequency >0.005 and common to both datasets were extracted. To ensure the datasets could be jointly meta-analysed, these SNPs were checked for same effect alleles and flipped accordingly in R (https://www.r-project.org/).

### The meta-analysis method

MetaUSAT (Unified Score-based Association Test) is a software package for performing GWAS meta-analysis studies on genetically correlated phenotypes. (21) (https://github.com/RayDebashree/metaUSAT) MetaUSAT applies a multivariate meta-analysis approach instead of a univariate approach of analysing each related trait separately. Unlike traditional GWAS meta-analysis on a single trait, where several sets of summary statistics on a single trait are combined into a single summary measure for that trait, the multivariate meta-analysis implemented by metaUSAT does not combine the summary statistics; instead, a joint analysis is performed using summary statistics from related traits. It is a statistical inference approach that leverages related traits to provide a *P* value for the test of no association of any trait with a SNP against the alternative that at least one trait is associated with the SNP. Being a complex data-adaptive approach, metaUSAT does not output an overall effect size (Beta) and standard error (SE) values for each SNP. MetaUSAT is robust to the association structure of correlated traits and to potential sample overlap. (21)

### The annotation method

The output generated from metaUSAT was uploaded to FUMA v1.3.6b for SNP annotation. (22) FUMA also generates a Manhattan plot and a corresponding Q-Q plot for the meta-analysis result (https://fuma.ctglab.nl/). FUMA uses “maximum distance of linkage disequilibrium (LD) blocks to merge” (default value = 250kb) to determine the number of associated loci and the r^2^ value (default value r^2^ > 0.6 to be considered as non-independent) to determine the number of independent significant SNPs. In addition, the gene-based association analysis and the gene-set analysis were performed with MAGMA v1.08, which was integrated in FUMA. (23) In gene-based association analysis, summary statistics of SNPs were aggregated to the level of whole genes, testing the joint association of all SNPs in the gene with the phenotype. In other words, all the SNPs were mapped to 19,436 protein coding genes if the SNPs are located within genes.

In gene-set analysis, individual genes were aggregated to groups of genes sharing certain biological, functional or other characteristics. This was done to provide insight into the involvement of specific biological pathways or cellular functions in the genetic aetiology of a phenotype. A total of 10,894 gene sets were tested and a competitive test model was applied. Tissue expression analysis was obtained from GTEx (https://www.gtexportal.org/home/) which was also integrated in FUMA. In the tissue expression analysis, average gene-expression per tissue type was used as gene covariate to test positive relationships between gene expression in a specific tissue type and genetic associations. In addition, regional plots of the identified new loci were generated by LozusZoom. (http://locuszoom.org/)

## Results

There were 8,500,802 common SNPs from both cohorts analysed by the metaUSAT software. FUMA reported 38 independent genetic loci across autosomal chromosomes with the *LRP1-STAT6-SDR9C7* region in chromosome 12q13.3 being the most significantly associated locus with a leading *P* value of 1.24 × 10^−62^ for rs11172113. The *FHL5-UFL1* locus in chromosome 6q16.1 was the second most significantly associated, with a *P* value of 6.57 × 10^−39^ for rs9486715. A Manhattan plot showing these loci is shown in Figure 1. A corresponding Q-Q plot is included as Supplementary Figure 1. Among the 38 identified loci, there were 7,952 SNPs that demonstrated an association with genome-wide significance, with *P* value < 5 × 10^−8^. Among these SNPs, 133 SNPs were considered as independent associations (r^2^ < 0.6 with any SNP within the 7,952 SNPs). (Supplementary Table 1 and 2)

**Figure 1:**
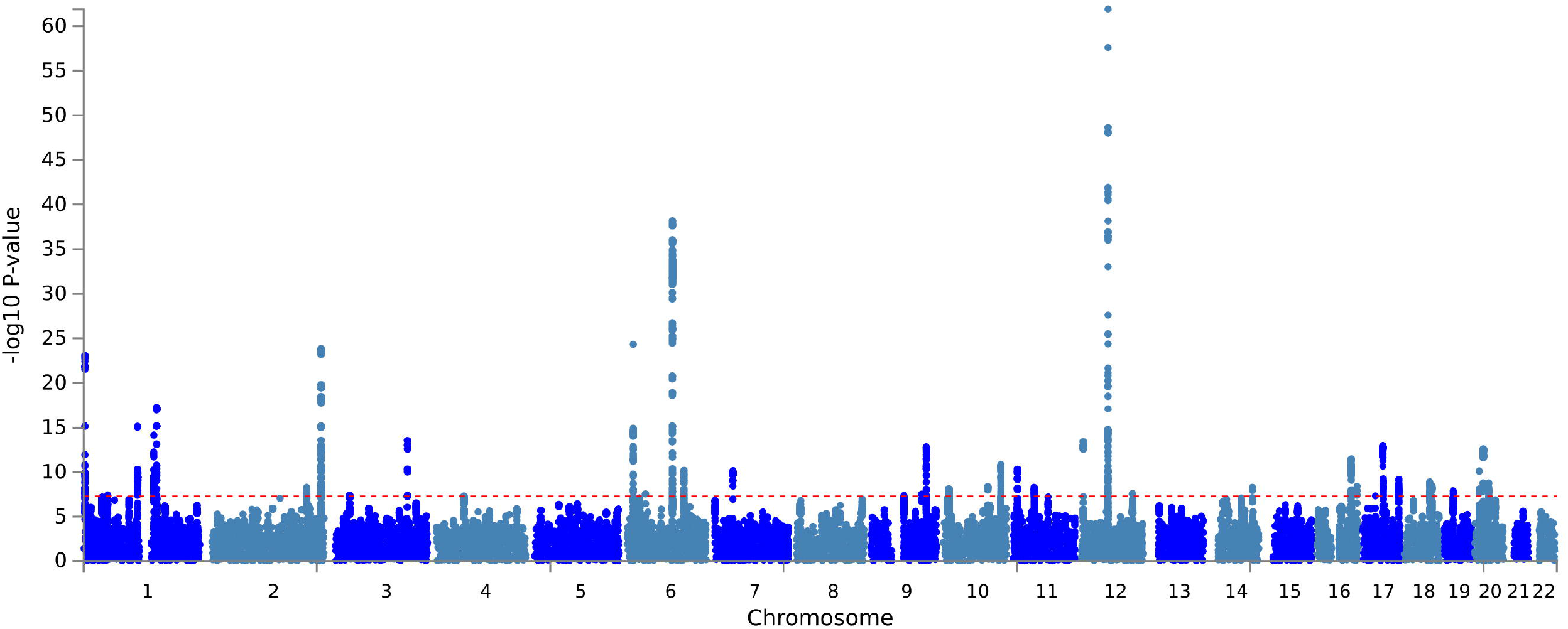
The Manhattan plot of the GWAS meta-analysis on headaches (N□ =□ 397,385). The dashed red line indicates the cut-off *P* value of 5□ × □10^−8^.

Table 1 summarises the information relating to the 38 associated loci. Among these loci, 25 loci had previously been reported by Gormley et al and 9 further loci had been separately identified by Meng et al (14,15). Four of the 38 loci were newly identified. The *ONECUT2* gene locus (18q21.31) was the strongest signal among these 4 new loci, associated with a *P* value of 1.29 × 10^−9^ for rs673939. Table 2 summarises the details of the 4 newly identified loci. Regional plots of these 4 new loci are included as Figure 2.

**Table 1:**
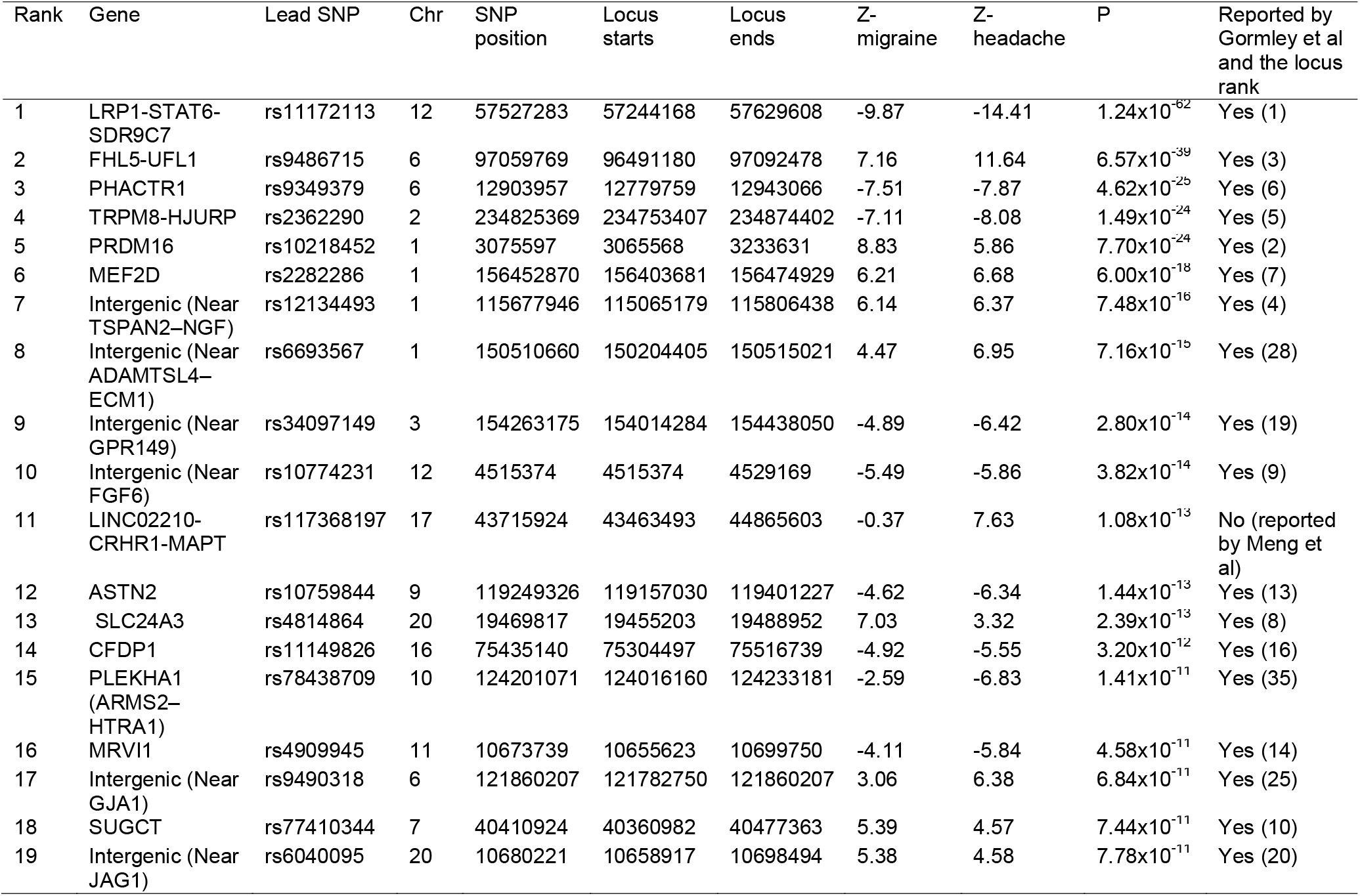
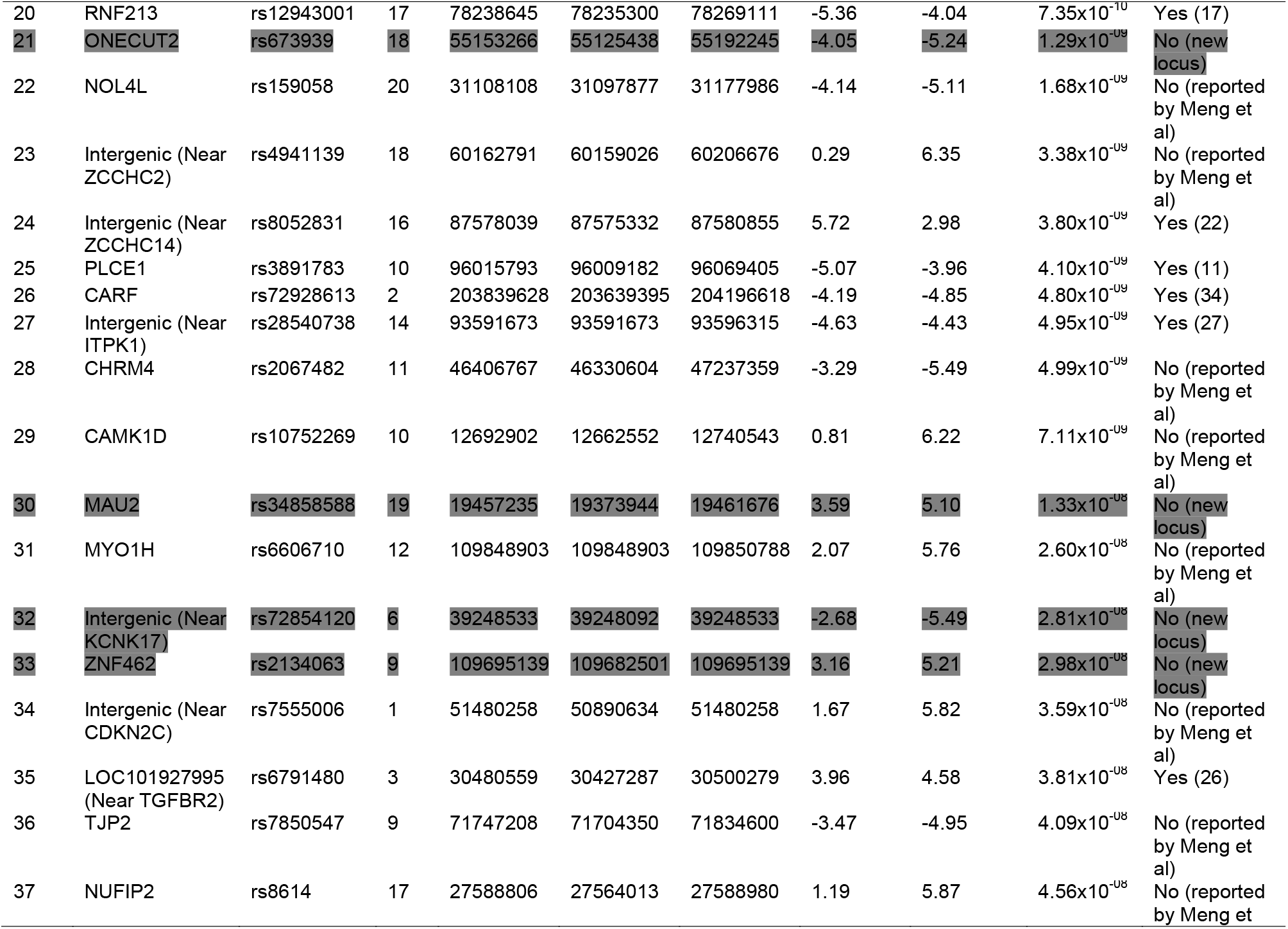
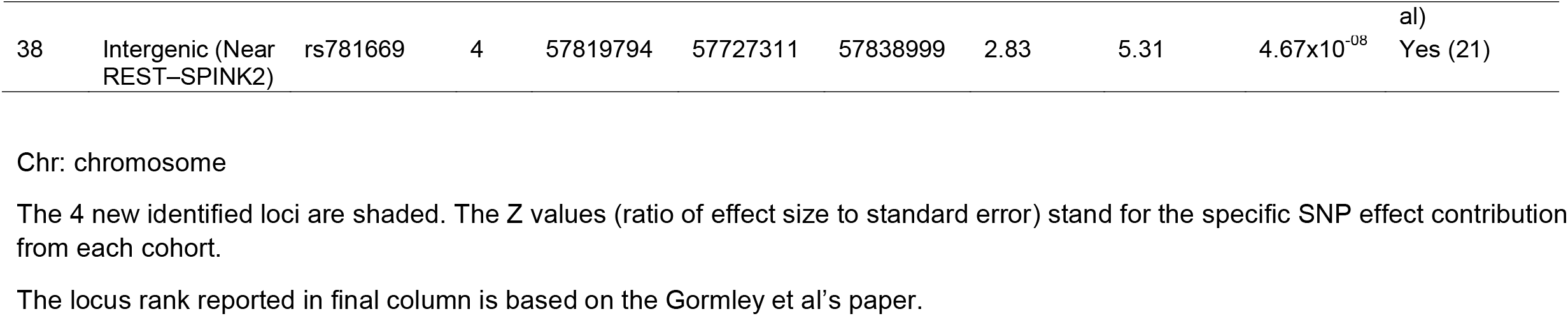
The 38 loci generated by the GWAS meta-analysis study on self-reported headache and self-reported migraine

**Table 2:**
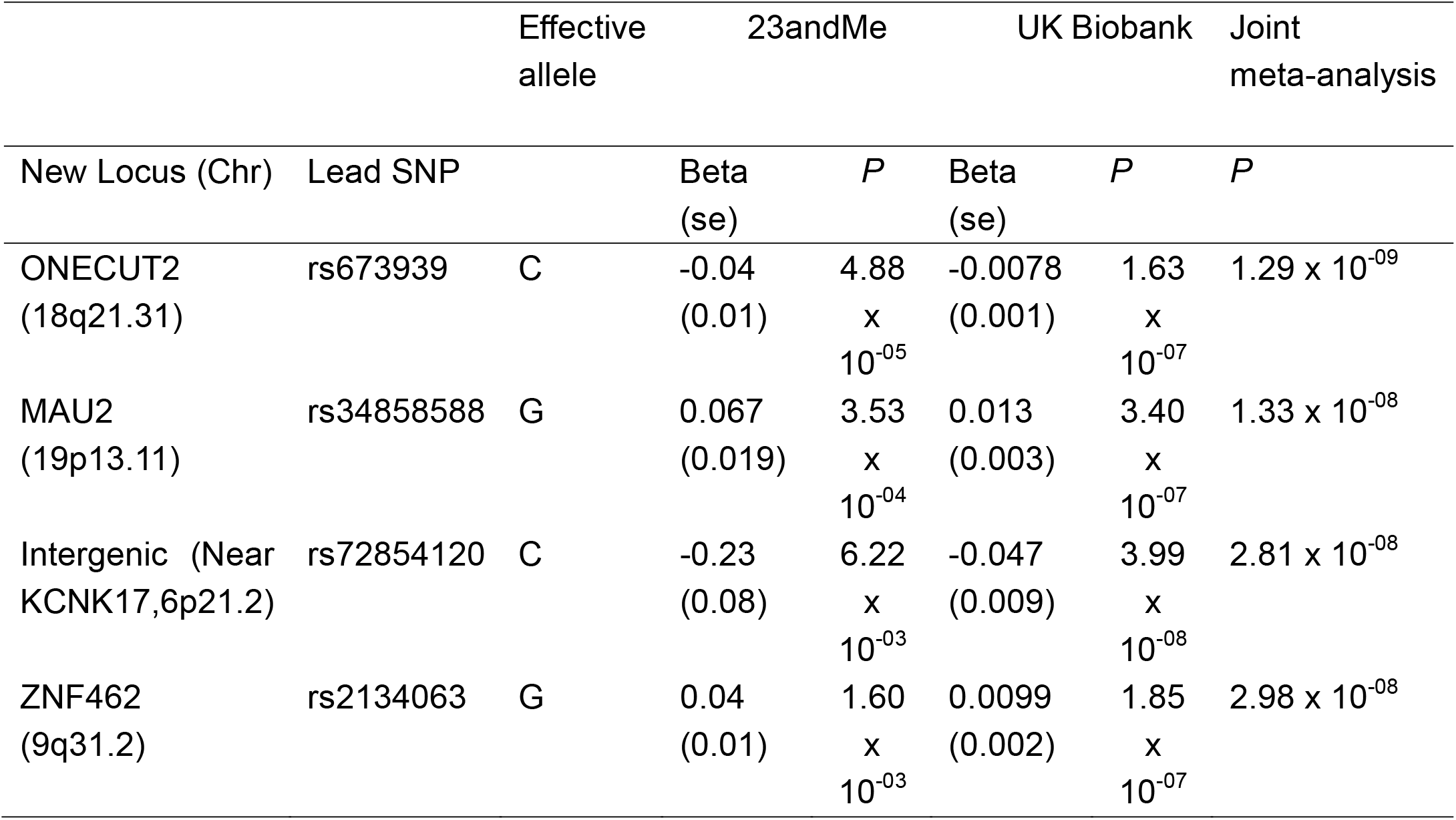
The summary statistics of the 4 new loci of headaches

**Figure 2:**
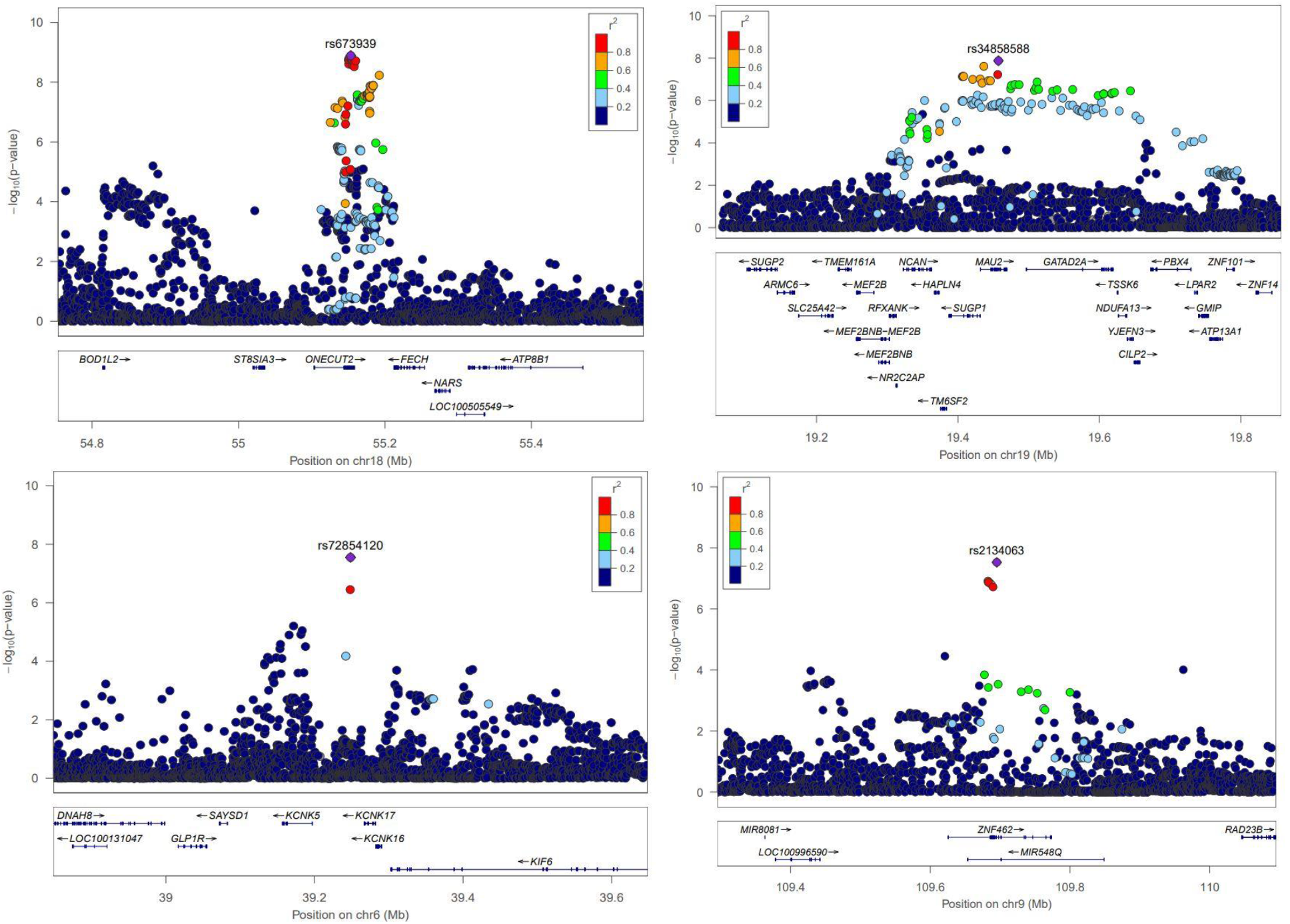
The regional plots of the 4 new loci. Up left: the ONECUT2 region; Up right: the MAU2 region; Bottom left: the Intergenic region (Near KCNK17,6p21.2); Bottom right: the ZNF462 region.

The gene-based association study identified 51 genes (cut-off *P* value = 0.05/19,436 = 2.57 × 10^−6^) that were associated with headaches, with the *PRDM16* gene showing the strongest association (*P* value of 7.10 × 10^−15^). All significantly associated genes are summarised in Supplementary Table 3.

The gene-set analysis found that no specific pathway was significantly associated with headaches after Bonferroni correction (cut-off *P* value = 0.05/10,894 = 4.6 × 10^−6^). The top 10 pathways are included in the Supplementary Table 4.

Two types of tissue analysis were performed. The tissue expression analysis on 30 general tissues revealed that both brain tissues and vascular tissues are potentially involved in the disease mechanisms (Figure 3). The tissue expression analysis on 54 specific tissues also found 11 brain tissues with significant association (*P* < 0.05/54 = 9.26 x10^−4^) (Figure 4).

**Figure 3:**
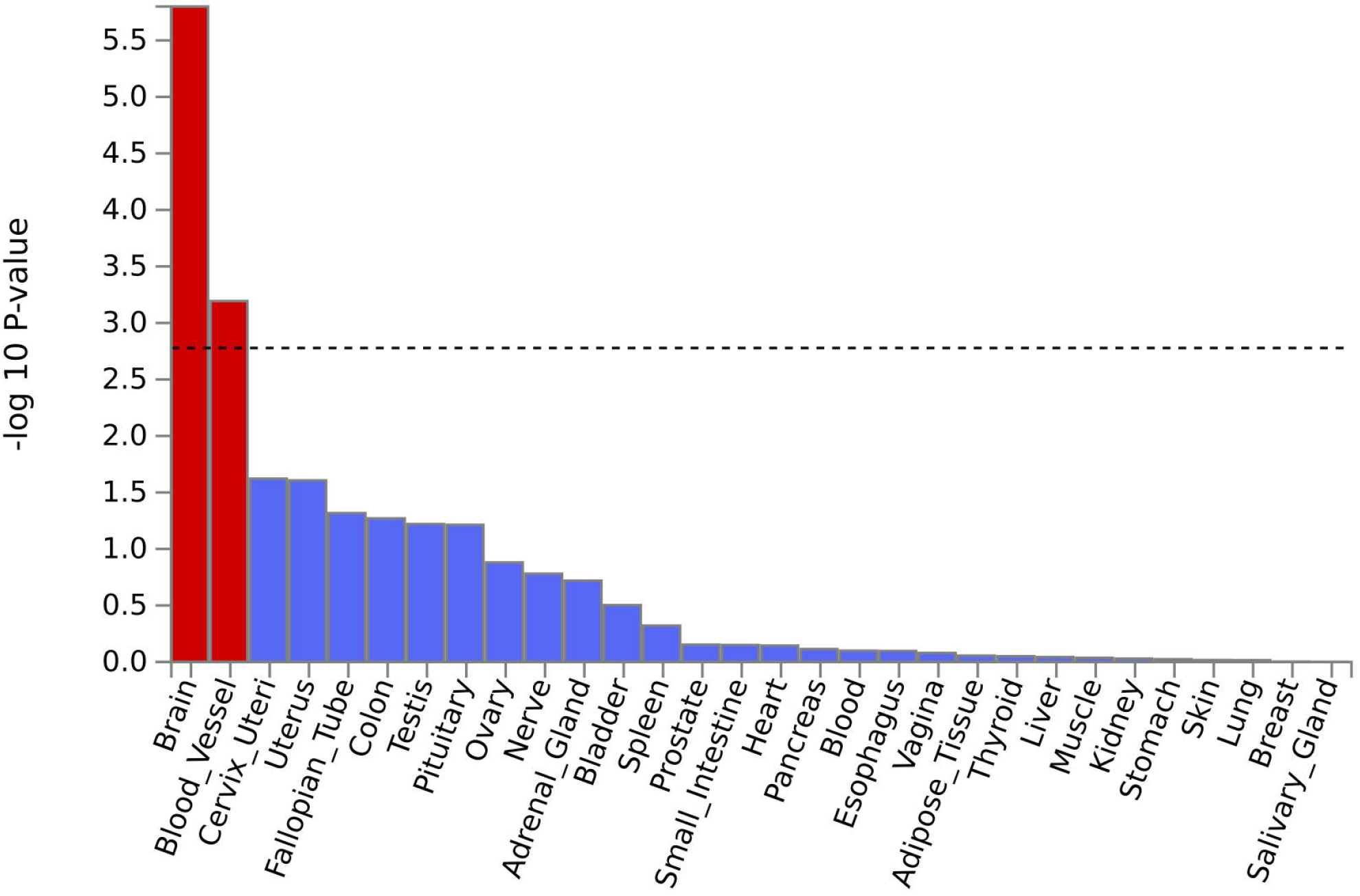
Tissue expression results on 30 specific tissue types by GTEx in the FUMA. The dashed line shows the cut-off P value for significance with Bonferroni adjustment for multiple hypothesis testing.

**Figure 4:**
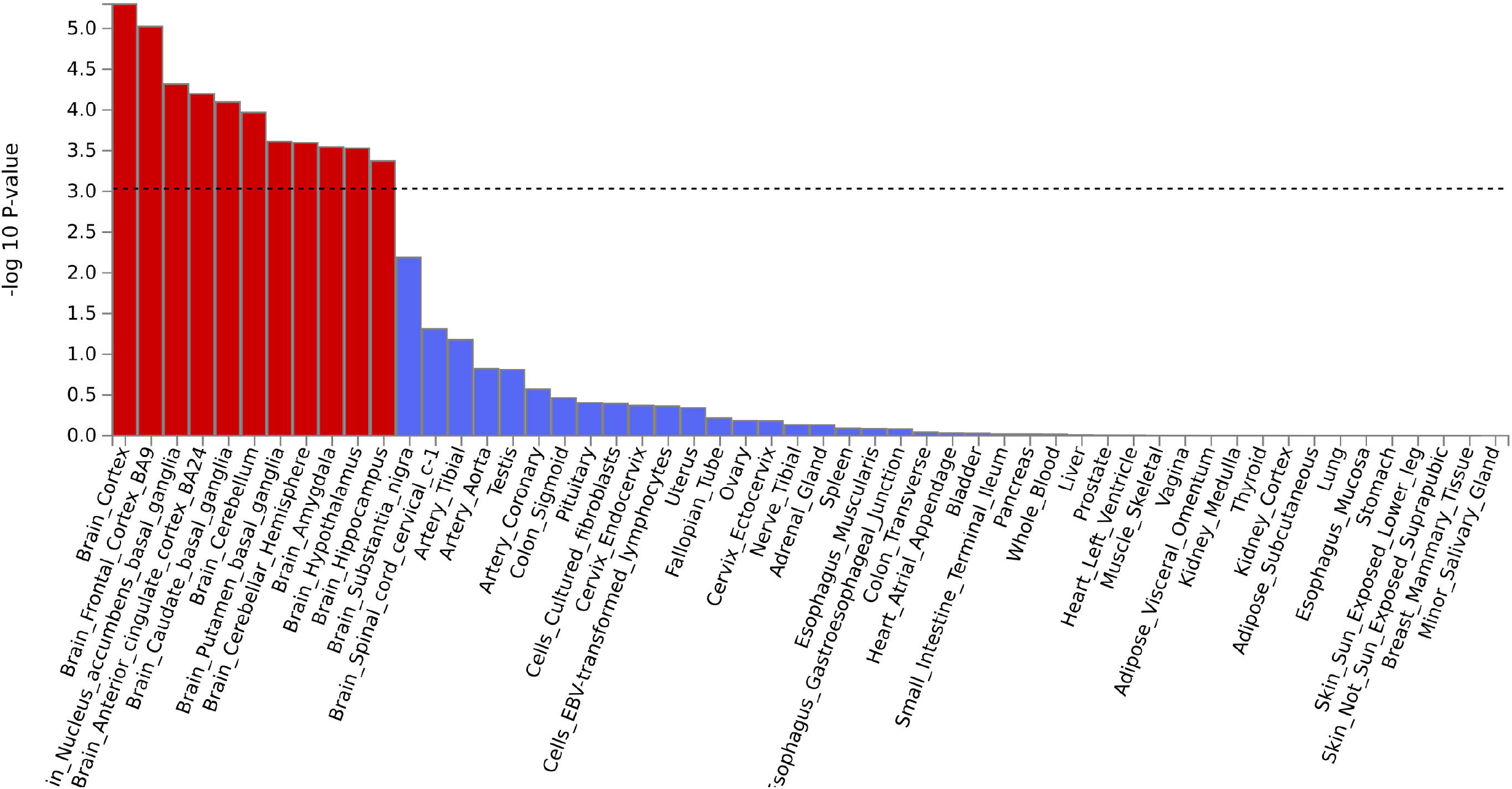
Tissue expression results on 53 specific tissue types by GTEx in the FUMA. The dashed line shows the cut-off P value for significance with Bonferroni adjustment for multiple hypothesis testing.

## Discussion

We performed a GWAS meta-analysis study on two highly genetically correlated phenotypes based on summary statistics from two large GWAS: self-reported headache and self-reported migraine (genetic correlation value = 0.72). This analysis identified 38 loci associated with headaches, of which 34 had been previously identified (9, 10) and 4 were newly identified loci.

GWAS on complex traits have achieved great success in the past decade. (24) Furthermore, GWAS meta-analysis on same phenotypes from multi-centers and multi-cohorts also improve statistical power to identify genetic variants which otherwise cannot be detected by a single cohort study. (25) However, the number of cohorts in a genetic consortium will reach a bottleneck when most of the existing cohorts are already included. It becomes more difficult to include extra cohorts into the consortium to achieve a higher study power. Meanwhile, it might also be challenging to fund and allocate resources to increase sample size of cohorts in a genetic consortium. This has led to the development and use of statistical methods that leverage other aspects of a study to increase detection power. Software developed for joint meta-analysis of GWAS on existing correlated phenotypes can improve power with minimum additional resource requirement, particularly as it is now a routine requirement for GWAS summary statistics to be shared publicly after publication. (26). Specific software created for this purpose includes META-SCOPA and metaCCA. (27,28)

In this study, we used a recently developed software called metaUSAT, whose properties have been illustrated using simulated data, and the T2D-GENES and the METSIM datasets.(29,30) The software does not generate an overall Beta or standard error for each SNP but it calculates a Z value representing the contribution of each SNP from each cohort or each phenotype. As with Beta values, if Z values are both positive or both negative, then the direction of the SNP effect on the traits is the same. In Table 2, we see that the most strongly associated SNPs in the top nine loci all have the same direction of effect. However, the lead SNP in the *LINC02210-CRHR1-MAPT* locus (ranked 10^th^), although highly significantly associated, showed a different direction of effect in the two datasets. This might indicate that the role of this locus might be different in these two phenotypes. Comparing the Z-values could be a novel way to differentiate the genetic impact of certain SNPs in genetically correlated yet different phenotypes. (31)

Consistent with previous studies, the most significantly associated locus in the *LRP1-STAT6-SDR9C7* region was the strongest locus identified in the meta-analysis (*P* = 1.24 × 10^−62^ for rs11172113). (9,10) This locus, ranging from 57,244,168 to 57,629,608 in chromosome 12, contained 122 SNPs associated with genome-wide significance, among the 166 SNPs in the output dataset. The *LRP1* gene has been well established as a migraine gene. (11, 12) One theory about its possible link with migraine is that the LRP1 protein interacts with the glutamate receptors on neurons while the pathophysiology of migraine has been suggested to be related with the glutamate homeostasis. (32) The gene-based association study revealed that the *PRDM16* gene was the most significantly associated gene, followed by *CRHR1, MAPT*, and *KANSL1* (Supplementary Table 3). Through the tissue expression analysis, both brain and vascular tissues were indicated as being involved in the mechanisms of headaches. Gormley et al found that vascular factors played a main role in migraine (9), while in our UK Biobank study, we found that neural tissues were major factors in self-reported headache. We therefore deduce that for other types of headaches, such as tension-type headache which produce most headaches in the general population, the role of neural tissue is likely to be greater than that of vascular factors.

In this study, we found 4 new loci which have not previously been reported to be associated with headaches. The *ONECUT2* gene region was the most strongly associated among these 4 loci. The Z values from the headache study (Z = -5.24) and the migraine study (Z = -4.05) were in the same direction. The original *P* values of the top SNP (rs673939) in this region were 9.00 × 10^−8^ in the Meng et al study (10) and 4.88 x10^−5^ in the 23andMe migraine dataset (9). The *ONECUT2* gene, also termed *OC⍰2*, is a newly discovered member of the ONECUT transcription factor family. (33) *ONECUT2* can widely regulate the protein expression associated with cell proliferation, migration, adhesion and differentiation, thus being involved in the regulation of the development of an organism. (34) It has been well reported for its associations with multiple cancers. Although we do not know why it is statistically associated with headaches, the gene is expressed in the brain. (https://www.ncbi.nlm.nih.gov/gene/9480) It is not uncommon that SNPs can be associated with multiple phenotypes which seem completely unrelated. (35) *MAU2* is a protein-coding gene, which plays an important role when cohesins (chromosome-associated multi-subunit protein complex) try to bind to DNA to carry out a large spectrum of chromatin-related functions, including sister chromatid cohesion, DNA repair, transcriptional regulation, and three-dimensional organization of chromatin. (36) Mutations of *MAU2* have been linked with a rare disorder of Cornelia de Lange Syndrome. (37) *KCNK17* is the nearest gene to the leading SNP of rs72854120 in the third new locus. Variants of this gene have been reported to be associated with ischaemic stroke, cerebral hemorrhage and arrhythmia. (38,39) The protein products of *ZNF462* have shown important roles in embryonic development in animal models. (40) Variants of this gene have been reported to contribute to craniofacial and neurodevelopmental abnormalities. (41) It is worth noting that the Z values of each leading SNP in the 4 new loci were all in the same direction in each of the two cohorts.

Although we were successful in performing this study, researchers need to be note that we had particular advantages in this study. One was that the two phenotypes are highly genetically correlated. If the two phenotypes are not so highly correlated, the ability of a study to detect more new variants would be reduced. Secondly, our two cohorts were both of mainly European descent and with minimum sample overlap; therefore, we avoided some negative impact (such as increasing type I and type II errors) which could be caused by these factors in the study.

It is also important to note that the control definitions in the two GWAS datasets were different. The controls used in the UK Biobank self-reported headache phenotype reported no pain within previous month, while the controls used in the 23andMe self-reported migraine phenotype could have had pain in body sites other than the head. This mean that genes identified in the UK Biobank cohort may not be specific to headache, but could be more generally associated with pain.

In summary, our study identified 4 new genetic loci which are associated with self-reported headaches and/or migraine, and shed further light on their potential mechanisms. Further research could attempt a meta-analysis study on GWAS of different types of primary headaches (on the condition that they are reasonably genetically correlated) to identify further genetic components.

## Data Availability

The GWAS meta-analysis summary statistics can be downloaded from https://app.box.com/s/gm2qkf17hc9w1fc5ymvifgbxz0httvhs
The FUMA results can be viewed from
https://fuma.ctglab.nl/browse/334.
Data from 23andMe were obtained under a data transfer agreement. Further information about obtaining access to the 23andMe Inc. summary statistics is available from: https://research.23andme.com/collaborate/. Any other data relevant to the study that are not included in the article or its supplementary materials are available from the authors upon reasonable request.

https://app.box.com/s/gm2qkf17hc9w1fc5ymvifgbxz0httvhs

## Funding

This work was supported by the STRADL project [Wellcome Trust, grant number: 104036/Z/14/Z]. The funders had no role in study design, data collection, data analysis, interpretation, writing of the report.

## Conflict of interest

The employees of 23andMe/23andMe Research Team hold stock in the company. The other authors declare that they have no conflict of interest.

## Acknowledgements

The current study was conducted under approved UK Biobank data application number 4844. We would like to thank all participants of the UK Biobank cohort who have provided necessary genetic and phenotypic information. We would like to thank the research participants and employees of 23andMe for making this work possible.

23andMe Consortium

The following members of the 23andMe Research Team contributed to this study: Stella Aslibekyan, Adam Auton, Elizabeth Babalola, Robert K. Bell, Jessica Bielenberg, Katarzyna Bryc, Emily Bullis, Daniella Coker, Gabriel Cuellar Partida, Devika Dhamija, Sayantan Das, Sarah L. Elson, Teresa Filshtein, Kipper Fletez-Brant, Pierre Fontanillas, Will Freyman, Anna Faaborg, Shirin T. Fuller, Pooja M. Gandhi, Karl Heilbron, Barry Hicks, Ethan M. Jewett, Katelyn Kukar, Keng-Han Lin, Maya Lowe, Jey C. McCreight, Matthew H. McIntyre, Steven J. Micheletti, Meghan E. Moreno, Joanna L. Mountain, Priyanka Nandakumar, Elizabeth S. Noblin, Jared O’Connell, Yunru Huang, Aaron A. Petrakovitz, Vanessa Lane, Aaron Petrakovitz, Joanne S. Kim, G. David Poznik, Morgan Schumacher, Anjali J. Shastri, Janie F. Shelton, Jingchunzi Shi, Suyash Shringarpure, Vinh Tran, Joyce Y. Tung, Xin Wang, Wei Wang, Catherine H. Weldon, Peter Wilton, Alejandro Hernandez, Corinna Wong, Christophe Toukam Tchakouté.

## Data availability

The GWAS meta-analysis summary statistics can be downloaded from https://app.box.com/s/gm2qkf17hc9w1fc5ymvifgbxz0httvhs

The FUMA results can be viewed from

https://fuma.ctglab.nl/browse/334.

Data from 23andMe were obtained under a data transfer agreement. Further information about obtaining access to the 23andMe Inc. summary statistics is available from: https://research.23andme.com/collaborate/. Any other data relevant to the study that are not included in the article or its supplementary materials are available from the authors upon reasonable request.

## Author contribution

W.M. organised project, drafted the paper and contributed to the analysis. P.S.R., C.N. and A.L.R. performed the meta-analysis. H.H. contributed to Table 1. M.J.A. performed the UK Biobank GWAS analysis. H.Z. and Z.H.L. contributed to discussion parts. The 23andMe Research Team provided the GWAS summary statistics of the 23andMe cohort. D.R. supervised the usage of the metaUSAT software and provided comments. L.C. and C.N.A.P. provided comments to the paper. A.M. and B.H.S. organised the project and provided comments.

## Notes

Conflicts of Interest: The employees of 23andMe/23andMe Research Team hold stock in the company. The other authors declare that they have no conflict of interest.

### Author Declarations

The current study was conducted under approved UK Biobank data application number 4844.

