## Supplementary figures and images for "A meta-analysis of the genome-wide association studies on two genetically correlated phenotypes (self-reported headache and self-reported migraine) identifies four new risk loci for headaches (N=397,385)"

### S figure 1

#### Slide 1
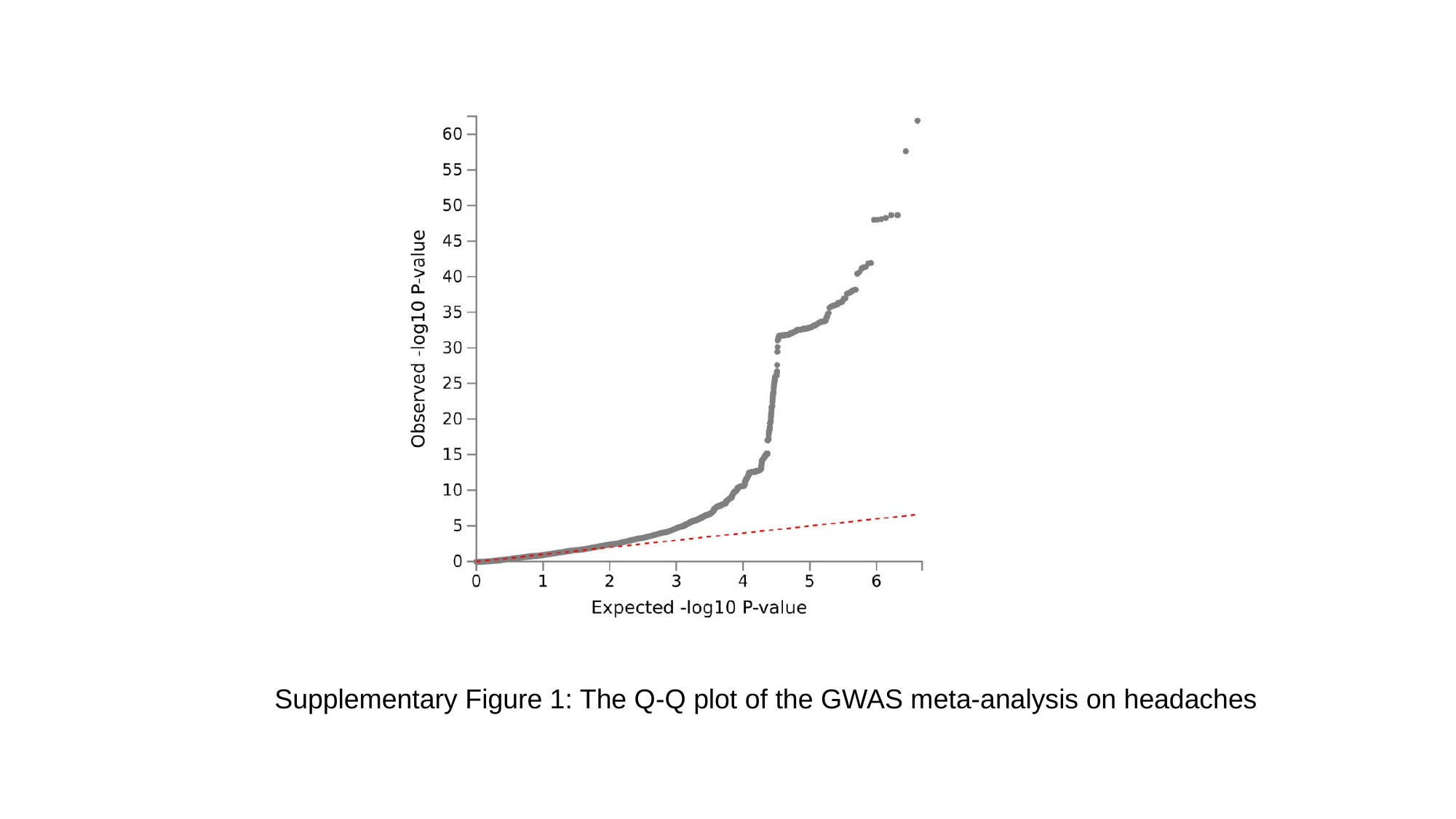

Supplementary Figure 1: The Q-Q plot of the GWAS meta-analysis on headaches
